# Investigating the causal nature of the relationship of subcortical brain volume with smoking and alcohol use

**DOI:** 10.1101/2020.09.03.20187385

**Authors:** Emma Logtenberg, Martin F Overbeek, Joëlle A Pasman, Abdel Abdellaoui, Maartje Luijten, Ruth J van Holst, Jacqueline M Vink, Damiaan Denys, Sarah E Medland, Karin JH Verweij, Jorien L Treur

**Author notes:** Shared first author (Emma Logtenberg, Martin F Overbeek).

## Abstract

**Background:** Structural variation in subcortical brain regions has been linked to substance use, including the most prevalent substances nicotine and alcohol. It may be that pre-existing differences in subcortical brain volume affect smoking and alcohol use, but there is also evidence that smoking and alcohol use can lead to structural changes. We assess the causal nature of this complex relationship with bi-directional Mendelian randomization (MR).

**Methods:** MR uses genetic variants predictive of a certain trait (‘exposure’) as instrumental variables to test causal effects on a certain outcome. Due to random assortment at meiosis, genetic variants shouldn’t be associated with confounders, allowing less biased causal inference. We employed summary-level data of the largest available genome-wide association studies of subcortical brain region volumes (nucleus accumbens, amygdala, caudate nucleus, hippocampus, pallidum, putamen, and thalamus; n = 50,290) and smoking and alcohol use (smoking initiation, n = 848,460; cigarettes per day, n = 216,590; smoking cessation, n = 378,249; alcohol drinks per week, n = 630,154; alcohol dependence, n = 46,568). The main analysis, inverse-variance weighted regression, was verified by a wide range of sensitivity methods.

**Results:** There was strong evidence that alcohol dependence decreased amygdala and hippocampal volume and that smoking more cigarettes per day decreased hippocampal volume. From subcortical brain volumes to substance use, there was no or weak evidence for causal effects.

**Conclusions:** Our findings suggest that heavy alcohol use and smoking can causally reduce subcortical brain volume. This adds to accumulating evidence that alcohol and smoking affect the brain, and most likely mental health, warranting more recognition in public health efforts.

## Introduction

Subcortical brain regions have consistently been implicated in substance use, playing a crucial role in the brain’s reward system^1^. It is thought that addiction reflects a vicious cycle of intoxication, withdrawal and craving, with (subcortical) brain circuits mediating these three stages^2^. The causal nature of the relationship between structural variation in subcortical brain regions and substance use is largely unclear. Subcortical brain volume and substance use are both substantially heritable, and there is evidence that they share (part of) their genetic aetiology^3^. Alternatively, the relationship may be causal, such that pre-existing differences in subcortical brain volume assert a direct effect on substance use. Causal effects in the other direction are also plausible, i.e., substance use affecting brain structure. Most likely, the complex relationship between subcortical brain volume and substance use is due to a combination of these mechanisms, making it challenging to identify causal effects.

Nicotine and alcohol are responsible for the majority of substance use related morbidity and mortality worldwide^4,5^. Most literature on the relationship of subcortical brain volumes with smoking and alcohol use is based on relatively small cross-sectional studies, reporting mixed findings. Smoking has been associated with smaller nucleus accumbens^6,7^, amygdala^7–9^, hippocampus^10^, pallidum^9^ and thalamus^7,9,11^ volumes, to smaller^8^ and larger^12^ caudate volume, and to larger putamen volume^6^. Alcohol (ab)use has been associated with smaller nucleus accumbens^13,14^, amygdala^15^, hippocampus^13,14,16^, pallidum^14,17^ and with smaller^18^ and larger^19^ caudate, smaller^13,14,16^ and larger^20^ thalamus and smaller^14^ and larger^20^ putamen volumes. Recently, the ENIGMA addiction working group attempted to resolve these inconsistent findings with a mega-analysis of subcortical thickness and surface area (volume being its product), among 1,628 controls and 2,277 individuals with dependence on alcohol, nicotine, cocaine, methamphetamine, and/or cannabis^21^. Smoking was associated with greater thickness and surface area for all regions, with the strongest associations for the nucleus accumbens and the hippocampus, while alcohol dependence was associated with lower thickness and surface area, with the strongest associations for the hippocampus, amygdala, thalamus, and putamen.

Few longitudinal imaging studies have investigated relationships between subcortical brain volume and substance use. Recently, a study was published that obtained structural brain measures and extensive survey-data for 714 individuals at ages 14 and 19. Using a machine learning method, the authors found that alcohol and cannabis use were associated with accelerated cortical thinning and a (mild) increase in subcortical volumes^22^. While these were longitudinal analyses, the study’s observational nature means there is potential for bias due to (unmeasured) confounding and reverse causality. Whereas a randomized controlled trial (RCT) to assess causal relationships between subcortical brain volume and substance use would be unfeasible, a promising alternative is Mendelian randomization (MR)^23^. Instead of experimental manipulation, MR uses genetic variants as proxies for the proposed independent variable. Because genes are randomly transmitted from parents to offspring at conception, genetic variants should not be associated with confounders (e.g. socio-economic status). Reverse causation is also not possible, as the genetic independent variable is fixed at birth.

MR has been applied to assess the relationship of smoking and alcohol use with psychiatric disorders. For instance, there was evidence that ADHD increases smoking and alcohol dependence and also that smoking causally increases ADHD risk^24^. Bi-directional causal relationships were also reported between smoking and depression^25^, while depression was found to causally increase alcohol dependence risk without evidence for the reverse^26^. Individuals with psychiatric disorders, including ADHD^27^ and depression^28^, are known to have smaller subcortical volumes compared to healthy controls. It may be that causal effects between smoking/alcohol and psychiatric disorders are mediated by subcortical brain volume. A recent MR study found no evidence for a causal effect of smoking on hippocampal volume^29^, but the analyses were based on much smaller samples than currently available and other subcortical regions were not included. We conduct the first comprehensive MR study using the largest genetic data-sets available of the volume of seven subcortical brain regions (nucleus accumbens, amygdala, caudate nucleus, hippocampus, pallidum, putamen, thalamus) and substance use (smoking initiation, cigarettes smoked per day, smoking cessation, alcohol drinks per week, and alcohol dependence), to probe bi-directional, causal relationships.

## Methods and materials

### Mendelian randomization

Mendelian randomization (MR) is based on the premise that genetic markers can be used as proxies for a variable that is hypothesized to be a risk factor, or ‘exposure’, for another ‘outcome’ variable. Single Nucleotide Polymorphisms (SNPs) are the most commonly used genetic markers. The validity of MR relies on three core assumptions: 1) the association of the genetic instrument with the exposure is robust (ensured by selecting SNPs that reached genome-wide significance, i.e. *p*<5e–08); 2) the instrument is not associated with confounding variables; 3) the instrument does not influence the outcome through any other path than the exposure. Horizontal pleiotropy, where a SNP directly affects multiple traits, could lead to the second and third assumptions being violated. In order to assess whether the assumptions were met, we applied a range of sensitivity methods, described below.

### Data

We took summary-level data from a published GWAS on subcortical brain volumes (n = 13,171^30^) and meta-analyzed these with summary-level data from a GWAS in 37,119 UK Biobank participants (**Supplementary Methods**). This resulted in a total sample of 50,290 for the volume of the nucleus accumbens, amygdala, caudate nucleus, hippocampus, pallidum, putamen, and thalamus. For substance use, we used summary-level data from the single largest available GWAS on smoking and alcohol use^31^ (smoking initiation n = 848,460, cigarettes per day n = 216,590, smoking cessation n = 378,249, drinks per week n = 630,154; note that UK Biobank was excluded to prevent sample overlap and we meta-analyzed data from the remaining cohorts with data from 23andme, **Supplementary Methods**) and a separate GWAS on alcohol dependence^32^ (n = 46,568). The meta-analyses were n-weighted, due to measurement variance in the original samples, resulting in z scores. To allow MR analysis, we constructed beta coefficients and standard errors using these z scores, the effect allele frequencies and sample size^33^. While the unit of MR estimates based on such constructed betas and standard errors cannot be reliably interpreted, the direction of effect and statistical strength of the evidence can.

Because we obtained the exposure estimates and the outcome estimates from separate samples, it’s impossible to verify if individuals in the outcome sample were affected by said exposure. Therefore, when we refer to an exposure causally affecting an outcome, this should be interpreted as an effect of the ‘liability to’ that exposure.

### Main analysis

For clarifying purposes, we describe the main analysis approach for one specific relationship, where hippocampal volume is the exposure (i.e., the independent variable) and smoking initiation is the outcome (i.e., the dependent variable). First, SNPs that robustly predict hippocampal volume (*p*<5E–08) were identified in the hippocampus GWAS, and their effect size estimates and standard errors extracted. These SNPs – i.e., the genetic instrument – were then identified in the smoking initiation GWAS, and their effect size estimates and standard errors extracted. To estimate the causal effect, the SNP-smoking initiation association was divided by the SNP-hippocampal volume association for each individual SNP, and the estimates of multiple SNPs combined with Inverse-Variance Weighted (IVW) regression. IVW provides the first indication of a causal effect by indicating the degree to which SNPs that predict the exposure (hippocampal volume), also predict the outcome (smoking initiation). We tested causal relationships with subcortical brain volumes as the exposures and smoking initiation, cigarettes per day, smoking cessation, alcohol per week and alcohol dependence as the outcomes – and in the other direction, with smoking initiation, alcohol per week and alcohol dependence as the exposures and subcortical volumes as the outcomes. If less than 10 SNPs *p*<5e–08 were available, we additionally constructed a genetic instrument containing SNPs under a more lenient p-value threshold of *p*<1e–05. We clumped SNPs for independence at *r*^2^< 0.01 and 10,000 kb^34^.

Because the GWAS for cigarettes per day consisted of smokers only^31^, SNPs from that study aren’t appropriate to use as proxies for cigarettes per day in never smokers. Therefore, the complete subcortical brain volume dataset (n = 50,290), consisting of both smokers and never smokers, could not be used. For UK Biobank participants (n = 37,119), we had information on smoking behaviour available and could perform GWASs of subcortical brain volumes in never smokers (n = 22,555) and in ever smokers (n = 14,564). We then applied summary-level MR with cigarettes per day as the exposure, in never and ever smokers separately. This approach provides an additional test of horizontal pleiotropy, as it allowed us to check MR assumptions 2 and 3 – the genetic instrument should not be associated with the outcome through other routes than the exposure. If the genetic instrument for cigarettes per day predicts subcortical brain volume in never smokers, this indicates horizontal pleiotropy because there can’t be a true causal effect^35^.

### Sensitivity analyses

The F-statistic was computed to assess instrument strength for all exposures – F>10 reflecting a sufficiently strong instrument^36^. In order to test the robustness of an IVW finding, we applied six sensitivity methods with different and partly contrasting assumptions. First, weighted median regression, which produces a reliable causal estimate if 50% or more of the total weight of the genetic instrument comes from valid (not biased) SNPs^37^. Second, weighted mode regression, which clusters the SNPs in the genetic instrument based on their causal estimates, and selects the estimate of the SNP-cluster with the largest weight as the final causal estimate. This results in an unbiased value if the SNPs in that cluster are valid and the most common causal effect estimate is indeed the true causal effect^38^. Third, MR-Egger, which permits the intercept to deviate from zero, allowing a formal test of horizontal pleiotropy (when there is no horizontal pleiotropy, the intercept is zero)^39^. MR-Egger is reliable if the InSIDE (Instrument Strength Independent of Direct Effect) assumption is met, meaning that the strength of the instrument (SNP-exposure association) should not correlate with the direct effect of the SNPs on the outcome. MR-Egger also requires sufficiently strong genetic instruments, indicated as the NOME (No Measurement Error) assumption. This can be assessed with the IGX^2^ statistic, ranging between 0 and 1. A lower value represents a higher chance that the NOME assumption is violated^40^. If IGX^2^ is ≥0.9, NOME is unlikely to be violated and the results can be reliably interpreted. If IGX^2^ is 0.6 – 0.9, NOME may have been violated but this can be corrected with MR-Egger simulation extrapolation (SIMEX). If IGX^2^ is < 0.6, MR-Egger results are likely biased and can’t be reliably interpreted. Fourth, GSMR (Generalised Summary-data-based Mendelian Randomization), which accounts for very low levels of linkage disequilibrium (LD) between SNPs and sampling variance in the estimated SNP effects, to attain higher statistical power. GSMR identifies and removes SNPs that are likely outliers based on their effect size (HEIDI-filtering)^41^. Fifth, MR-PRESSO (Pleiotropy Residual Sum and Outlier), which compares the observed residual sum of squares to the expected residual sum of squares for each SNP, and re-runs outlier-corrected IVW analyses^42^. Sixth, Steiger filtering, which identifies potential bias from reverse causation. It calculates the amount of variance that each SNP explains in both the exposure and the outcome and tests whether the explained variance is, as would be expected, higher for the exposure than the outcome. SNPs that explain more variance in the outcome than the exposure are excluded^43^.

In addition, we computed Cochran’s Q statistic to assess heterogeneity across the causal estimates of the SNPs included in each instrument^36^ – high heterogeneity points to horizontal pleiotropy. It should be noted that it is also possible for a true causal effect to run through multiple, very separate biological pathways, resulting in heterogeneity. To assess variability in the power of the genetic instruments, we computed the amount of variance that each instrument explained in the proposed exposure variable^44^.

Analyses were conducted using the TwoSampleMR package for R^34^, the GSMR package for R^41^ and the MR-PRESSO package for R^42^.

### Appraisal of the evidence

We didn’t correct for multiple testing explicitly because we analyse phenotypes for which, a priori, there are plausible hypotheses why they are (causally) associated and we want to avoid appraising the evidence based on an arbitrary threshold. We ascribe a finding as showing strong evidence, evidence, weak evidence, or no clear evidence for a causal effect, based on both the IVW regression – adhering to the interpretation of p-values suggested by Sterne and Davey Smith (2001)^45^ – and the sensitivity methods. Because sensitivity methods rely on stricter assumptions than IVW their statistical power is lower. It is to be expected that the statistical evidence, but not the effect size, decreases with stricter sensitivity methods, even for a true causal effect.

## Results

All genetic instruments showed sufficient strength as indicated by the mean F-statistic, ranging between 15.23 and 68.94 (**Table S1**). Based on the IGX^2^ statistic, MR-Egger could reliably be performed for all relationships, except when smoking initiation was the exposure (**Tables S2 and S3**). The amount of variance that the genetic instruments for substance use explained in the corresponding substance use variables ranged between 0.56% and 1.43%. For subcortical brain volumes it ranged between 0.17% and 3.97% (**Table S4**). A graphical display of all relationships with (weak or strong) evidence for causality is provided in **Figure 1**.

**Figure 1.**
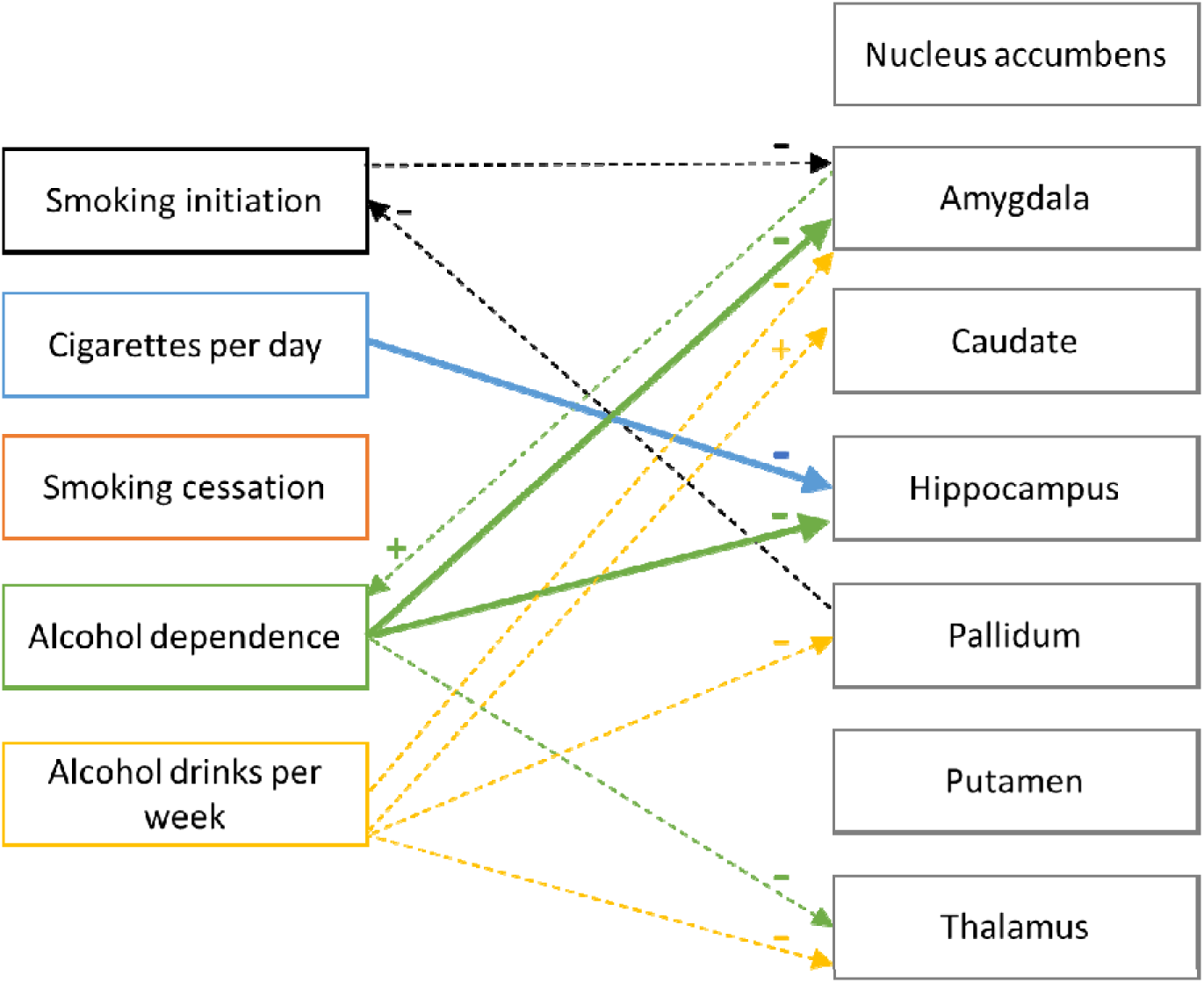
Graphical display of the relationships for which there was evidence for causality. Minus (–) signifies a negative, decreasing effect, while plus (+) signifies a positive, increasing effect. The thicker lines reflect evidence or strong evidence while the dotted, thinner lines signify weak evidence for causality. Note that for all relationships causal effects were tested in both directions, except for smoking cessation which was only tested as an outcome variable.

### Causal relationships from subcortical volumes to substance use

There was weak evidence that a larger pallidum volume decreased the odds of initiating smoking (beta_IVW_ = –0.04, *p* = 0.053). Weighted median, weighted mode, and GSMR corroborated this finding, showing similar effect sizes and stronger statistical evidence (**Table 1**). While there was no clear evidence for horizontal pleiotropy (Egger-intercept = –0.003, *p* = 0.332; **Table S5**), the regression coefficient of MR-Egger did not indicate a causal effect (**Table 1**). There was strong evidence for heterogeneity among the SNP-effects (Cochran’s Q *p* = 2.4E–05; **Table S6**). MR-PRESSO identified 2 SNP-outliers but there was no distortion of the causal estimate after outlier removal (**Table S7**). Steiger filtering did not identify SNPs that explained more variance in the outcome than the exposure (**Table S8**).

**Table 1.**
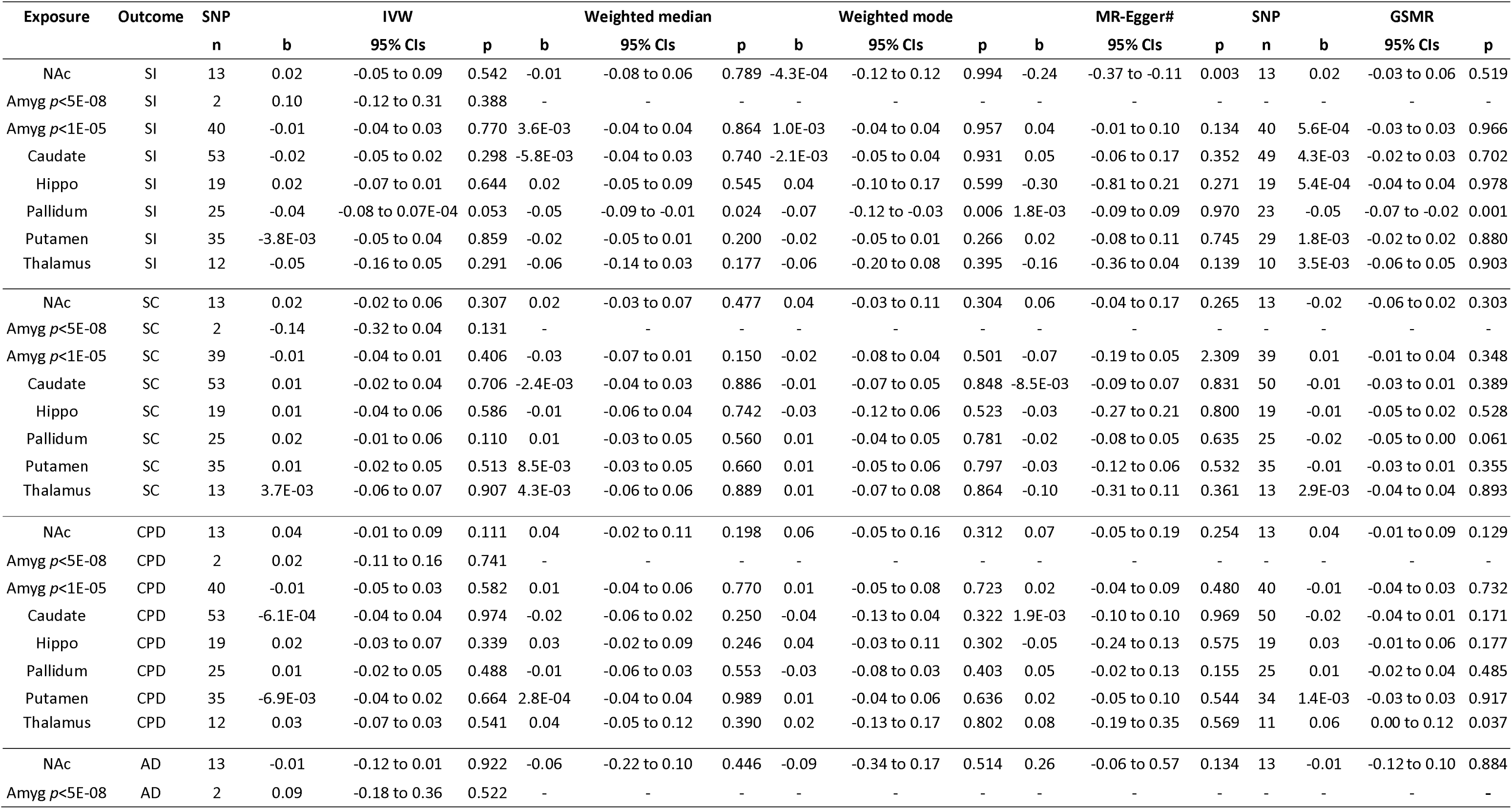
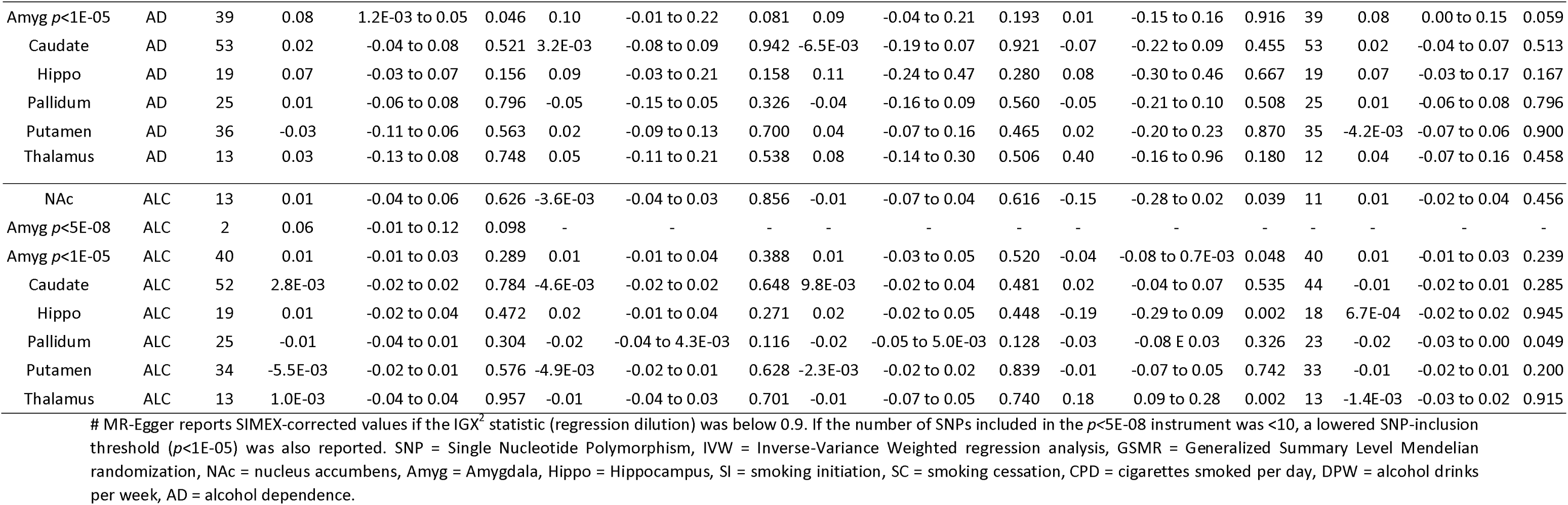
Mendelian randomization analysis with subcortical brain volumes as the exposures and smoking and alcohol use as the outcomes.

There was weak evidence that a larger amygdala volume increased alcohol dependence risk (*p*< 1E-05 beta_IVW_ = 0.08, *p* = 0.046), corroborated by weighted median, weighted mode and GSMR sensitivity methods, but not MR-Egger. There was no clear evidence for horizontal pleiotropy (Egger-intercept = 0.003, *p* = 0.400), nor for heterogeneity (*p* = 0.621). MR-PRESSO did not identify any SNP- outliers nor did Steiger filtering identify SNPs that explained more variance in the outcome than the exposure. With a 2-SNP instrument (*p*< 5E-08), there was a similar sized, positive effect, but no clear statistical evidence (beta_IVW_ = 0.09, *p* = 0.522).

There was very weak evidence that a larger amygdala volume increased the number of alcohol drinks per week (beta_IVW_ = 0.06, *p* = 0.098), but sensitivity analyses were not possible due to the *p*<5E–08 instrument only containing 2 SNPs, and with 40 SNPs under *p*<1E–05 there was no clear evidence for an effect (beta_IVW_ = 0.01, *p* = 0.289).

None of the other analyses showed clear evidence for causal effects of subcortical volumes on substance use.

### Causal relationships from substance use to subcortical volumes

There was weak evidence that smoking initiation decreased amygdala volume (beta_IVW_ = –0.05, *p* = 0.046), but the effect was only consistent with the GSMR method (**Table 2**). There was no clear evidence for horizontal pleiotropy (Egger-Intercept = –0.001 *p* = 0.457), but strong evidence for heterogeneity (Cochran’s Q *p* = 2.4E-07). MR-PRESSO identified 1 SNP-outlier, which did not impact the results. Steiger filtering excluded 44 SNPs but after running the analyses with the 302 remaining SNPs evidence for a causal effect remained (beta_IVW_ = –0.06, *p* = 0.013).

**Table 2.** Mendelian randomization analysis with smoking and alcohol use as the exposures and subcortical brain volumes as the outcomes.

| Exposure | Outcome | SNP | IVW |  |  | Weighted median |  |  | Weighted mode |  |  | MR-Egger# |  | SNP |  | GSMR |  |  |
| --- | --- | --- | --- | --- | --- | --- | --- | --- | --- | --- | --- | --- | --- | --- | --- | --- | --- | --- |
|  |  | n | b | 95% CIs | p | b | 95% CIs | p | b | 95% CI | p | b | 95% CIs | p | n | b | 95% CIs | p |
| SI | NAc | 346 | -0.04 | -0.10 to 0.01 | 0.114 | -0.03 | -0.10 to 0.04 | 0.381 | -0.02 | -0.13 to 0.09 | 0.743 | - | - | - | 339 | -0.05 | -0.09 to 0.00 | 0.050 |
| SI | Amygdala | 346 | -0.05 | -0.07 to -9.3E-04 | 0.046 | 4.0E-04 | -0.07 to 0.07 | 0.991 | 0.01 | -0.11 to 0.09 | 0.856 | - | - | - | 340 | -0.05 | -0.09 to 0.00 | 0.058 |
| SI | Caudate | 346 | 3.33E-03 | -0.10 to 0.07 | 0.919 | -0.02 | -0.10 to 0.06 | 0.660 | -6.0E-03 | -0.14 to 0.13 | 0.929 | - | - | - | 335 | -0.01 | -0.06 to 0.04 | 0.671 |
| SI | Hippo | 346 | -2.20E-03 | -0.09 to 0.05 | 0.939 | -0.02 | -0.09 to 0.06 | 0.680 | 6.51E-04 | -0.13 to 0.13 | 0.992 | - | - | - | 340 | -0.02 | -0.07 to 0.03 | 0.449 |
| SI | Pallidum | 346 | -0.04 | -0.09 to 0.02 | 0.166 | -0.02 | -0.09 to 0.05 | 0.502 | -0.01 | -0.14 to 0.13 | 0.934 | - | - | - | 338 | -0.03 | -0.08 to 0.02 | 0.206 |
| SI | Putamen | 346 | 0.02 | -0.09 to 0.08 | 0.614 | -0.01 | -0.09 to 0.06 | 0.728 | -0.03 | -0.15 to 0.09 | 0.618 | - | - | - | 336 | 0.01 | -0.04 to 0.06 | 0.614 |
| SI | Thalamus | 345 | -0.03 | -0.09 to 0.03 | 0.304 | -0.02 | -0.09 to 0.06 | 0.640 | -0.01 | -0.13 to 0.12 | 0.900 | - | - | - | 340 | -0.01 | -0.06 to 0.04 | 0.666 |
| AD | NAc | 10 | -0.06 | -0.21 to 0.05 | 0.301 | -0.06 | -0.21 to 0.08 | 0.388 | -0.06 | -0.26 to 0.13 | 0.545 | -0.32 | -0.77 to 0.12 | 0.189 | 10 | -0.07 | -0.20 to 0.06 | 0.309 |
| AD | Amygdala | 10 | -0.15 | -0.31 to -0.04 | 0.007 | -0.17 | -0.31 to -0.04 | 0.013 | -0.17 | -0.34 to -0.01 | 0.074 | -0.28 | -0.60 to 0.05 | 0.276 | 10 | -0.15 | -0.28 to -0.02 | 0.023 |
| AD | Caudate | 10 | 0.04 | -0.14 to 0.16 | 0.552 | 2.1E-03 | -0.14 to 0.14 | 0.977 | 0.01 | -0.17 to 0.18 | 0.934 | 0.49 | -0.02 to 1.01 | 0.099 | 10 | 0.02 | -0.11 to 0.15 | 0.734 |
| AD | Hippo | 10 | -0.11 | -0.26 to -0.01 | 0.037 | -0.11 | -0.26 to 0.04 | 0.156 | -0.08 | -0.28 to 0.11 | 0.417 | -0.50 | -0.87 to -0.13 | 0.031 | 10 | -0.12 | -0.25 to 0.01 | 0.077 |
| AD | Pallidum | 10 | -0.02 | -0.13 to 0.11 | 0.756 | 0.03 | -0.13 to 0.18 | 0.737 | 0.12 | -0.07 to 0.32 | 0.252 | -0.50 | -1.08 to 0.08 | 0.130 | 10 | 0.01 | -0.12 to 0.14 | 0.883 |
| AD | Putamen | 10 | -0.03 | -0.16 to 0.08 | 0.624 | -0.01 | -0.16 to 0.13 | 0.874 | -0.02 | -0.21 to 0.16 | 0.806 | 0.13 | -0.37 to 0.62 | 0.630 | 10 | -0.01 | -0.14 to 0.12 | 0.891 |
| AD | Thalamus | 10 | -0.09 | -0.21 to 0.02 | 0.097 | -0.06 | -0.21 to 0.09 | 0.443 | -0.03 | -0.21 to 0.15 | 0.774 | -0.46 | -0.91 to -3.7E-03 | 0.084 | 10 | -0.01 | -0.14 to 0.12 | 0.891 |
| ALC | NAc | 92 | -0.13 | -0.39 to 0.04 | 0.122 | -0.18 | -0.39 to 0.04 | 0.111 | -0.17 | -0.38 to 0.05 | 0.137 | -0.18 | -0.46 to 0.10 | 0.200 | 86 | -0.13 | -0.26 to 0.01 | 0.070 |
| ALC | Amygdala | 92 | -0.11 | -0.46 to 0.04 | 0.147 | -0.25 | -0.46 to -0.03 | 0.025 | -0.24 | -0.48 to -2.1E-04 | 0.053 | -0.17 | -0.42 to 0.07 | 0.174 | 87 | -0.13 | -0.27 to 0.00 | 0.057 |
| ALC | Caudate | 92 | 0.20 | -0.06 to 0.39 | 0.032 | 0.18 | -0.06 to 0.41 | 0.138 | 0.09 | -0.12 to 0.31 | 0.401 | 0.14 | -0.16 to 0.45 | 0.361 | 85 | 0.14 | 0.00 to 0.28 | 0.054 |
| ALC | Hippo | 92 | -4.67E-03 | -0.34 to 0.14 | 0.949 | -0.14 | -0.34 to 0.06 | 0.170 | -0.18 | -0.40 to 0.03 | 0.100 | -0.10 | -0.33 to 0.14 | 0.411 | 87 | -0.03 | -0.17 to 0.10 | 0.635 |
| ALC | Pallidum | 92 | -0.15 | -0.52 to 0.03 | 0.096 | -0.29 | -0.52 to -0.05 | 0.017 | -0.31 | -0.53 to -0.09 | 0.007 | -0.39 | -0.68 to -0.10 | 0.010 | 87 | -0.17 | -0.31 to -0.04 | 0.013 |
| ALC | Putamen | 92 | -0.01 | -0.29 to 0.19 | 0.896 | -0.06 | -0.29 to 0.17 | 0.605 | -0.13 | -0.36 to 0.10 | 0.258 | -0.12 | -0.45 to 0.21 | 0.482 | 83 | -0.01 | -0.15 to 0.13 | 0.839 |
| ALC | Thalamus | 92 | -0.13 | -0.48 to 0.03 | 0.104 | -0.26 | -0.48 to -0.04 | 0.022 | -0.27 | -0.50 to -0.04 | 0.025 | -0.21 | -0.46 to 0.05 | 0.118 | 86 | -0.13 | -0.27 to 0.00 | 0.058 |

In the analyses stratified for smoking status, there was strong evidence that smoking more cigarettes per day decreased hippocampal volume in smokers (beta_IVW_ = –94.73, *p* = 1.8E-06; **Table 3**). Results were consistent with weighted median, weighted mode, MR-Egger and GSMR methods, albeit with a smaller effect size for the latter. There was no clear evidence for horizontal pleiotropy (Egger-intercept = 0.633, *p* = 0.568) nor heterogeneity (*p* = 0.357). No SNP-outliers were identified with MR-PRESSO. Steiger filtering identified 9 SNPs that were more predictive of the outcome than the exposure, but after excluding these (leaving 40 SNPs) strong evidence for causality remained, consistent across sensitivity methods. There was also weak evidence for a negative effect of cigarettes smoked per day on hippocampal volume in never smokers – indicating horizontal pleiotropy – with a much smaller, less significant effect size (beta_IVW_ = –30.40, *p* = 0.050) and less consistency across sensitivity methods. Taken together, some horizontal pleiotropy exists, but on top of that, there is likely a decreasing effect of cigarettes smoked per day on hippocampal volume.

**Table 3.**
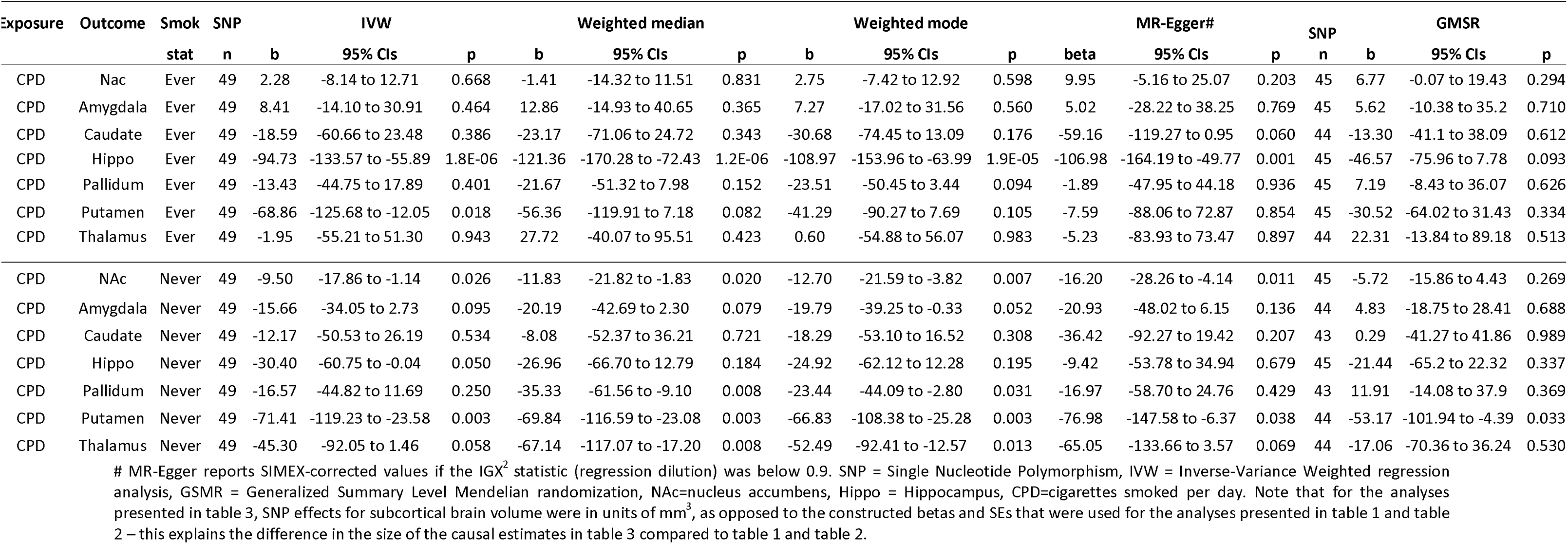
Mendelian randomization analysis with cigarettes per day as the exposure and subcortical brain volumes as the outcomes – stratified on smoking status (ever versus never smokers)

There was evidence for a negative effect of cigarettes smoked per day on putamen volume, but this relationship seems pleiotropic, given that the effect size and statistical evidence in ever and never smokers are nearly indiscernible (beta_IVW_ = –68.86, *p* = 0.018 and beta_IVW_ = –71.41, *p* = 0.003, respectively). Similarly, there was weak evidence for an effect of cigarettes per day on amygdala and thalamus volume in never smokers, pointing to horizontal pleiotropy.

There was strong evidence for a decreasing effect of liability to alcohol dependence on amygdala volume (beta_IVW_ = –0.15, *p* = 0.007) and evidence for a decreasing effect on hippocampal volume (beta_IVW_ = –0.11, *p* = 0.037). These results were consistent across weighted median, weighted mode, MR-Egger, and GSMR methods (**Table 2**). There was no clear evidence for horizontal pleiotropy for alcohol dependence-to-amygdala (Egger-intercept = 0.007, *p* = 0.468) and weak evidence for alcohol dependence-to-hippocampus (Egger-intercept = 0.023, *p* = 0.080). There was no clear evidence for heterogeneity in the SNP-effects (*p* = 0.950 and *p* = 0.691, respectively). MR-PRESSO did not identify SNP-outliers nor did Steiger filtering exclude SNPs that explain more variance in the outcome than the exposure.

There was weak evidence that alcohol dependence decreased thalamus volume (beta_IVW_ = – 0.09, *p* = 0.097), which was corroborated by MR-Egger, but not the other sensitivity methods. There was no clear evidence for horizontal pleiotropy (Egger-intercept = 0.021, *p* = 0.150), nor was there heterogeneity (*p* = 0.493). No SNP-outliers were identified with MR-PRESSO and Steiger filtering did not exclude any SNPs.

There was evidence that more alcoholic drinks per week increased caudate volume(beta_IVW_ = 0.20, *p* = 0.032). The effect was consistent with weighted median, but attenuated with weighted mode, MR-Egger and GSMR. There was no clear evidence for horizontal pleiotropy (Egger-intercept = 0.001, *p* = 0.629), but strong evidence for heterogeneity (*p* = 6.4E–10). MR-PRESSO identified 5 SNP-outliers, which did not distort the causal estimate. Steiger filtering identified 26 SNPs that explained more variance in the outcome than in the exposure and after removing these (leaving 66 SNPs) no evidence for a causal effect remained. Taken together, the evidence that alcoholic drinks per week has a positive effect on caudate volume was weak.

There was weak evidence that more drinks per week decreased pallidum volume (beta_IVW_ = – 0.15, *p* = 0.096), an effect which was consistent and even stronger in size and statistical evidence across weighted median, weighted mode, MR-Egger and GSMR. There was evidence for horizontal pleiotropy (Egger-intercept = 0.003, *p* = 0.049) and strong evidence for heterogeneity (*p* = 2.2E-08). MR-PRESSO identified 2 SNP-outliers, but there was no distortion in the causal estimate before and after outlier removal. With Steiger filtering 35 SNPs were excluded (leaving 58 SNPs), but after running the analyses again weak evidence for a causal effect remained.

Finally, from drinks per week to amygdala and thalamus volume there were sizable negative effects which, while there was no clear evidence for the main IVW method, appeared much stronger with the different sensitivity methods (**Table 2**). There was no horizontal pleiotropy (Egger-intercept = 0.001, *p* = 0.537 and 0.001, *p* = 0.459, respectively) but there was evidence for heterogeneity (*p* = 0.005 and *p* = 0.001, respectively). There were no SNP-outliers with MR-PRESSO and while Steiger filtering excluded 27 and 25 SNPs, respectively, weak evidence for causality remained.

## Discussion

This is the first study that applied Mendelian randomization to assess bi-directional, causal relationships between the volume of subcortical brain regions and a range of substance use behaviours. Our most robust findings were that (liability to) alcohol dependence causally decreased amygdala and hippocampal volume, and smoking more cigarettes per day causally decreased hippocampal volume.

The evidence that alcohol dependence decreased amygdala and hippocampal volume was particularly strong. This is in line with work showing that in individuals with alcohol dependence subcortical brain regions are smaller and have a lower thickness and surface area than in healthy controls – with the largest differences reported for the amygdala and hippocampus^13–18,21,46^. Given MR’s powerful premise and the consistency of our findings across sensitivity analyses, we are able to make stronger conclusions that this is due to causal effects of alcohol. It had previously been hypothesised that alcohol can cause cell death or reduced cell density, subsequently resulting in volume loss^46^. For instance, chronic alcohol consumption is known to induce the release of tumour necrosis factor alpha (TNF-α), a cytokine involved in potentiating neuro-inflammation which can cause neuronal death^47^. We found only weak evidence that more alcoholic drinks per week decreases amygdala, pallidum, and thalamus volume. This discrepancy in strength of evidence is likely due to the fact that alcohol dependence is the more severe phenotype, reflecting prolonged and heavy exposure of the brain to alcohol.

We found strong evidence that smoking more cigarettes per day (in smokers) decreases hippocampal volume and weak evidence that being a smoker versus being a non-smoker decreases amygdala volume – implying that exposure to cigarette smoking can induce structural subcortical brain changes. While the literature on potential biological mechanisms responsible for such effects is scarce, animal work has shown that exposure to nicotine can induce apoptosis in hippocampal cells^48,49^. In contrary to our findings, and to those of other observational studies^6–11^, a large ENIGMA study found smoking to be associated with greater thickness and surface area of all subcortical regions^21^. This discrepancy may be due to the fact that the ENIGMA-study was observational and its findings influenced by confounding factors.

There is an ongoing discussion as to whether differences in brain structure between substance (ab)users and controls reflect pre-existing differences, or whether they are the result of alterations caused by substance use. Our results mostly point to the latter, with robust evidence for negative effects of alcohol dependence and smoking on some subcortical volumes, without (similarly robust) evidence for causal effects from subcortical volumes to substance use. This is important knowledge with potentially far reaching consequences. Volume loss might lead to cognitive deficits and a higher chance of developing mental illness, given that smaller volume of the amygdala and hippocampus has been implicated in the most common psychiatric disorders^50,51^. For instance, it is thought plausible that smoking-related structural brain changes in regions that connect fear response areas (e.g. amygdala) impact trait anxiety states, subsequently leading to an anxiety disorder^52^. More research is needed to explicitly test pathways from smoking and alcohol use to subcortical brain volume, and subsequently to psychiatric symptoms.

The current study has some important strengths. We used the largest available genetic datasets, which allowed us to test causal effects with sufficiently powered genetic instruments in both directions. We used a diverse and extensive set of sensitivity methods in order to assess the robustness of our findings and whether or not the assumptions underlying MR were met, allowing us to make claims about causality with considerable certainty. There are also limitations to note. While MR can provide less biased causal inference, there may be bias stemming from sources that have so far been less emphasised. One important source is ‘genetic nurturing’, which occurs when the genotype of parents directly affects offspring phenotypes even if the responsible genetic variants weren’t transmitted^53^. Second, assortative mating, i.e., spouses showing higher phenotypic similarity than expected by chance, may impact MR estimates if this similarity arises because individuals with a particular genetic predisposition choose their mate based on a genetically influenced phenotype^53^. The effect of both phenomena is that bias from confounding is reintroduced. Finally, while the GWASs we employed corrected for population structure, some geographic clustering may remain^54^. When these become available, large-scale within-family GWAS would be able to correct for more fine-grained (geographical/family) clusters, providing superior genetic estimates to use in MR^53^.

In sum, we report robust evidence that heavy alcohol use causally affects the brain, decreasing subcortical brain volume (at least as it pertains to the amygdala and the hippocampus). There was also considerable, but more tentative, evidence that smoking causally decreases amygdala and hippocampus volume. These findings provide additional proof that smoking and alcohol use can have a detrimental effect on the brain and it may implicate structural changes as a pathway connecting substance use to the development of (other) psychiatric disorders. We feel that, combined with accumulating evidence from other types of research, this justifies more recognition in public health efforts and clinical practice.

## Data Availability

The manuscript reports on analyses which were performed with publicly available summary-level genetic datasets

## Acknowledgements

JLT is supported by a Veni grant from the Netherlands Organization for Scientific Research (NWO; grant number 016.Veni.195.016). ML is also supported by a Veni grant from NWO (grant number 451–15–029). KJHV, AA, and JLT are supported by the Foundation Volksbond Rotterdam. AA is supported by ZonMw grant 849200011 from The Netherlands Organisation for Health Research and Development. SEM was supported by NHMRC grants APP1103623, APP1158127 and APP1172917. We acknowledge SURFsara for the usage of the Lisa cluster computer (supported by NWO, 15725).

